# SARS-CoV-2 Omicron variant: exploring healthcare workers’ awareness and perception of vaccine effectiveness: a national survey during the first week of WHO variant alert

**DOI:** 10.1101/2021.12.27.21268431

**Authors:** Mohamad-Hani Temsah, Fadi Aljamaan, Shuliweeh Alenezi, Khalid Alhasan, Abdulkarim Alrabiaah, Rasha Assiri, Rolan Bassrawi, Ali Alhaboob, Fatimah Alshahrani, Mohamad Alarabi, Ali Alaraj, Nasser S Alharbi, Rabih Halwani, Amr Jamal, Ayman Al-Eyadhy, Naif AbdulMajeed, Lina Alfarra, Wafa Almashdali, Fahad Al-Zamil, Mazin Barry, Ziad A. Memish, Jaffar A. Al-Tawfiq, Sarah Alsubaie

**Affiliations:** College of Medicine, King Saud University, Riyadh11362, Saudi Arabia; Pediatric Department, King Saud University Medical City, King Saud University, Riyadh11362, Saudi Arabia; Prince Abdullah Ben Khaled Celiac Disease Research Chair, Department of Pediatrics, Faculty of Medicine, King Saud University, Riyadh11362, Saudi Arabia; Critical Care Department, King Saud University Medical City, King Saud University, Riyadh11362, Saudi Arabia; Department of Psychiatry, College of Medicine, King Saud University Medical City, King Saud University, Riyadh11362, Saudi Arabia; Department of Basic Medical Sciences, College of Medicine, Princess Nourah bint Abdulrahman University, Riyadh 11564, Saudi Arabia; Division of Infectious Diseases, Department of Internal Medicine, King Saud University Medical City, King Saud University, Riyadh11362, Saudi Arabia; Department of Medicine, College of Medicine, Qassim University, Qassim51452, Saudi Arabia; Dr. Sulaiman Al Habib Medical Group, Riyadh11643, Saudi Arabia; Sharjah Institute of Medical Research, University of Sharjah, Sharjah27272, United Arab Emirates; Department of Clinical Sciences, College of Medicine, University of Sharjah, Sharjah 27272, United Arab Emirates; Department of Family and Community Medicine, King Saud University Medical City, Riyadh 11362, Saudi Arabia; Pediatric nephrology department, Prince Sultan Military Medical City, Riyadh11159, Saudi Arabia; Department of Ob-Gyn, Dr. Abdul Rahman Al Mishari Hospital, Riyadh12241, Saudi Arabia; Department of Ob-Gyn, Dr. Fatina Imran Medical Complex, Doha233, Qatar; Division of Infectious Diseases, Faculty of Medicine, University of Ottawa K1H 8M5, Canada; King Saud Medical City, Ministry of Health & Alfaisal University, Riyadh11533, Saudi Arabia; Hubert Department of Global Health, Emory University, Atlanta, Georgia30322, USA; Specialty Internal Medicine and Quality Department, Johns Hopkins Aramco Healthcare, Dhahra34465, Saudi Arabia; Infectious Disease Division, Department of Medicine, Indiana University School of Medicine, Indianapolis, IN46202, USA; Infectious Disease Division, Department of Medicine, Johns Hopkins University School of Medicine, Baltimore, MD21218, USA

**Keywords:** COVID-19 vaccine, SARS-CoV-2 Omicron variant, healthcare workers’ perceptions, SARS-CoV-2 variants

## Abstract

**Background:** As the SARS-CoV2 Omicron variant spreads in several countries, healthcare workers’ (HCWs) perceptions and worries regarding vaccine effectiveness and boosters warrant reassessment.

**Methods:** An online questionnaire among HCWs in Saudi Arabia (KSA) was distributed from Dec 1^st^ to 6^th^ 2021 to assess their perceptions, vaccine advocacy to the Omicron variant, and their perception of the effectivness of infection prevention measures and vaccination to prevent its spread, their Omicron variant related worries in comparison to the other variants, and their agreement with mandatory vaccination in general for adults.

**Results:** Among the 1285 HCW participants, two-thirds were female, 49.8 % were nurses, 46.4% were physicians, and 50.0% worked in tertiary care hospitals. 66.9% considered vaccination to be the most effective way to prevent the spread of the Omicron variant and future variants. The respondents however perceived social distancing (78.0%), universal masking (77.8%), and avoiding unnecessary travel (71.4%) as slightly superior to vaccination to prevent the spread of SARS-CoV-2 variants. HCWs aging 55 or older agreed singficanlty with vaccine ineffictivness to control Omicron spread, while those who believed in non-pharmacolgical infection prevention measures agreed signifcantly with vaccination for that purpose. Male HCWs had a significant agreement with mandatory vaccination of all eligible adult populations. On the other hand, unwilling HCWs to receive the vaccine had strong disagreements with mandatory vaccination.

**Conclusions:** The current study in the first week of Omicron **showed that** only two-thirds of HCWs felt that vaccination was the best option to prevent the spread of the **Omicron** variant, indicating the need for further motivation campaigns for vaccination and booster dose. HCWs had a strong belief in **infection prevention measures to contain the spread of SARS-CoV-2 variants** that should be encouraged and augmented.

## 1. Introduction

The B.1.1.529 variant of Severe Acute Respiratory Syndrome Coronavirus 2 (SARS-CoV-2) was first reported to the World Health Organization (WHO) from South Africa on November 26, 2021 [1]. This emerging variant was named Omicron and designated as a variant of concern (VOC). This variant has more than 30 mutations on its spike protein. And similar to the Alpha variant, it is associated with an S-gene target failure on a specific PCR assay due to the presence of 69–70 deletions [2]. Preliminary evidence suggests an increased risk of reinfection with this variant and high contagiousness [3,4]. The Omicron variant had been reported in 110 countries across all six WHO regions as of December 22, 2021[5]. [1,2]In addition, the B.1.1.529 variant was thought to have a higher transmissibility rate than other circulating variants [3,6,7]. More concern has arisen from the questionable whether current vaccines provide adequate protection against this variant [8]. A recent study showed a 41-fold decline of the geometric mean titer (GMT) inverse of the plasma dilution required for 50% reduction in infection (FRNT50) with Omicron compared to previous infection with the wild-type virus, with a three-fold reduction in FRNT50 with the mRNA vaccine BNT162b2[9]. However, it is likely that previous infection followed by vaccination, or a booster dose for those completing their primary series, will likely increase neutralization levels that would confer the needed protection against severe disease from the emerging Omicron variant [10]A recent study showed that even with Omicron, the rate of hospitalization was highest among unvaccinated and lowest among those who received a booster dose.88% likelihood to escape current vaccines[11-13].

As new variants emerge, HCWs must continue to serve COVID-19 and non-COVID patients while still fulfilling personal commitments for their families and themselves[14-16]. Burnout, emotional exhaustion, mental distress, depression and psychological stress have been linked to HCWs during this pandemic, and research continues to reveal high rates of infection [17]. In addition to patient care, HCWs must keep up with any developments in relation to emerging variants and their effects on clinical presentation, management, and measures of prevention—mainly vaccination and infection prevention practices[18,19]. As the Saudi Ministry of Health announced the first case of Omicron variant on Dec 1^st^, 2021[20], we undertook this study to assess HCWs’ awareness about the SARS-CoV2 Omicron variant, their worry levels in relation to the different SARS-CoV2 variants, their perception of the effectiveness of the current vaccines, and other preventive measures to inhibit the spread of the Omicron variant or other future variants, as well as to assess their agreement with mandatory vaccination of the adult population.

## 2. Method

### 2.1 Data Collection

This was a cross-sectional survey (Appendix 1) carried out among HCWs in Saudi Arabia (KSA), conducted between Dec 1 and 6, 2021. Participants were invited via a convenience sampling technique through various healthcare providers’ social media platforms, such as WhatsApp, Twitter, and emails.

HCWs were surveyed regarding their Omicron variant awareness, perceptions of the effectiveness of COVID-19 vaccinations and other measures to prevent its spread and that of other future variants, and their agreement with mandatory vaccination for all eligible adults. The last part of the survey focused on HCWs’ resilience, anxiety, and coping strategies, which was published as preprint [21]. The current survey was adopted from our previously published studies, with modifications related to the new Omicron variant [22-25]. The questionnaire was pilot-validated and sent through the SurveyMonkey© electronic platform as described previously [22-25].

Participants gave consent at the beginning of the survey, were notified about the purpose of the study, and participated voluntarily in the study. The Institutional Review Board at the College of Medicine and King Saud University approved the study (approval 21/01039/IRB).

### 2.2 Statistical analyses

The mean and standard deviation were used to assess continuous variables (worry level for example), frequency and percentage were used for categorically measured variables (agreement with mandatory vaccination for example, age was analyzed as categories). The 5-point Likert score was analyzed by categorical method to assess agreement versus disagreement, the participants’ responses were grouped into two groups: 1. Agree including agree and strongly agree responses; 2. Disagree including disagree, strongly disagree and neither agree nor disagree responses. A histogram and the K–S statistical test of normality were used to assess the statistical normality assumption of the continuous variables, and Levene’s test was used to assess the homogeneity of statistical variance assumption. A multiple-response dichotomies analysis was used to analyze the multiple response variables (participants’ source of information for example). Pearson’s correlations test (r) was used to assess the correlations between metric variables. Multivariate binary logistic regression analysis was used. The associations were expressed as the odds ratio and 95% confidence interval. The SPSS IBM statistical analysis program (Version#21. Armonk, NY: IBM Corp.) was used for the statistical data analysis. The statistical significance level (P-value) was considered at 0.01 if achieved, and 0.05 if 0.01 not achieved according to the used software.

## 3. Results

A total of 1285 HCWs completed the online survey. Table 1 shows the participants’ baseline characteristics. The majority (64%) were female, and 70.9% aged between 25 and 44 years. The majority were expatriates (62.3%). Regarding their clinical role, 49.8% were nurses and 46.4% were physicians, of whom 24% were consultants. Fifty percent worked in tertiary institutes, while two-thirds worked in the outpatient department (OPD) or general wards.

**Table.1:** Baseline characteristics of the participating HCWs. N=1285.

| Variable | Frequency | Percentage |
| --- | --- | --- |
| Sex |  |  |
| Female | 822 | 64.0 |
| Male | 463 | 36.0 |
| **Age Group** |  |  |
| 25-34 years | 434 | 33.8 |
| 35 to 44 years | 477 | 37.1 |
| 45-54 years | 273 | 21.2 |
| ≥ 55 years | 101 | 7.9 |
| **Nationality** |  |  |
| Saudi | 484 | 37.7 |
| Expatriate | 801 | 62.3 |
| **Clinical Role** |  |  |
| Consultant | 319 | 24.8 |
| Assistant Consultant/Fellow | 74 | 5.8 |
| Resident/Registrar | 203 | 15.8 |
| Nurse | 640 | 49.8 |
| Allied Health Practitioner | 49 | 3.8 |
| **Hospital Type** |  |  |
| Primary Healthcare Center | 338 | 26.3 |
| Secondary Hospital | 302 | 23.5 |
| Tertiary Hospital | 645 | 50.2 |
| **Hospital Working Area** |  |  |
| Intensive Care Unit (ICU) | 141 | 11.0 |
| Emergency Room (ER) | 91 | 7.1 |
| Operating Room (OR) | 41 | 3.2 |
| COVID-19 Isolation Ward | 53 | 4.1 |
| General Ward | 492 | 38.3 |
| Outpatient Department (OPD) | 368 | 28.6 |
| Non-Clinical Area | 99 | 7.7 |
| **Geographical Region** |  |  |
| Riyadh City and Central Region | 740 | 57.6 |
| Eastern Province | 71 | 5.5 |
| Western Province | 120 | 9.3 |
| Northern Province | 34 | 2.6 |
| Southern Province | 320 | 24.9 |

Almost all (97.9%) of the participants did not recently travel to countries where the Omicron variant has been recorded. Most (71%) HCWs had not been in contact with COVID-19 patients during the last 3 months, while 22.3% of the HCWs had developed COVID-19 previously.

Regarding COVID-19 vaccination, 99.5% had received two doses, and the first dose was divided equally between AstraZeneca ChAdOx1-S and Pfizer vaccines, and 75% of the second dose was of the Pfizer–BioNTech COVID-19 vaccine. Regarding the booster dose, 94.1% either received or planned to receive it once eligible, according to time-based criteria of the local regulations (Table 2).

**Table.2:** Descriptive analysis of the HCWs’ experiences of COVID-19 disease, screening and immunization.

| Variable | Frequency | Percentage |
| --- | --- | --- |
| **Have you been in contact with COVID-19 patients during the past 3 months?** |  |  |
| No | 912 | 71.0 |
| Yes | 373 | 29.0 |
| **Were you previously diagnosed with PCR-positive COVID-19 yourself?** |  |  |
| No | 999 | 77.7 |
| Yes | 286 | 22.3 |
| **Did you travel to any country where the Omicron variant has been recorded during the last month?** |  |  |
| Yes | 27 | 2.1 |
| No | 1258 | 97.9 |
| **Which vaccine did you receive for your first COVID-19 vaccine shot?** |  |  |
| AstraZeneca ChAdOx1-S | 600 | 46.7 |
| Moderna | 5 | 0.4 |
| Pfizer-BioNTech | 676 | 52.6 |
| Not received | 4 | 0.3 |
| **Which vaccine did you receive for your second COVID-19 vaccine shot?** |  |  |
| AstraZeneca ChAdOx1-S | 296 | 23.0 |
| Moderna | 20 | 1.6 |
| Pfizer-BioNTech | 963 | 74.9 |
| Not received | 6 | 0.5 |
| **Did you receive the third (booster) COVID-19 vaccine?** |  |  |
| Yes | 250 | 19.5 |
| No: not yet eligible for it | 566 | 44.0 |
| No: I do not want to receive it | 76 | 5.9 |
| No: but I am planning to register for it | 393 | 30.6 |

\* Mean (SD)

Table 3 shows the participating HCWs’ awareness and knowledge of the available literature concerning certain facts and queries related to the Omicron variant (key answers displayed in brackets). Almost all (93%) the participating HCWs knew that the Omicron variant was first described in South Africa; however, 81.7% believed that the Omicron variant was already reported in Saudi Arabia. Of the respondents, 66% knew that Omicron variant is more transmissible than the Delta variant, while 26.7% were not sure. The majority (71.8%) did not know that the Omicron variant spike protein has 22 mutations. Still, 56.2% expected that Omicron variant causes similar signs and symptoms to the original variant. Most (85.1%) of our cohort were aware that SARS-CoV-2 mutations are expected, while they were almost equally divided regarding their opinion on whether they were unsure or whether the Omicron variant may/may not cause more severe disease compared to previous variants. About half of the participants were unsure if mRNA or vector-based vaccines are effective against the Omicron variant. At least two-thirds of them were unsure if therapeutics such as monoclonal antibodies used against previous variants might be effective against the variant.

**Table 3:**
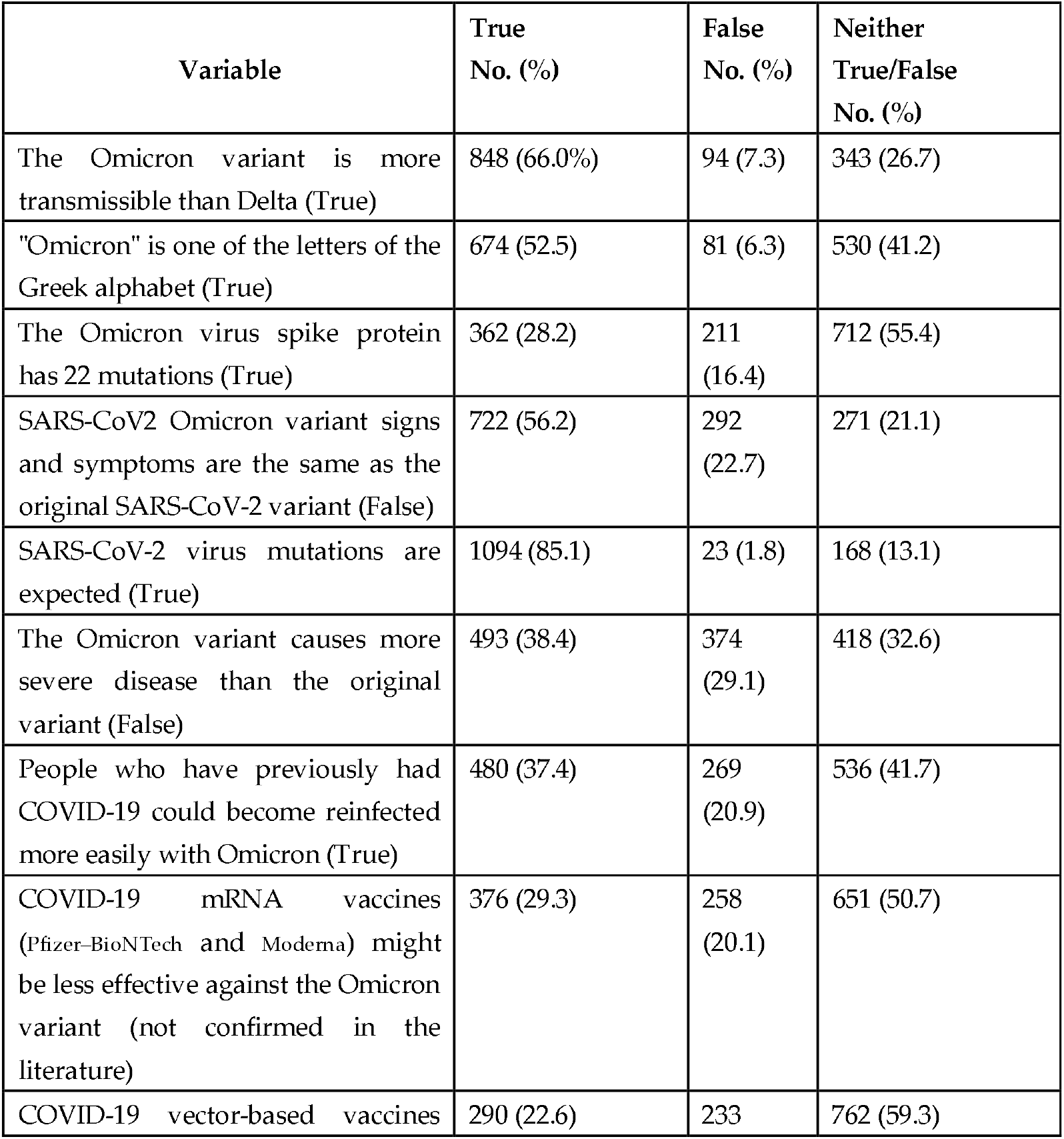
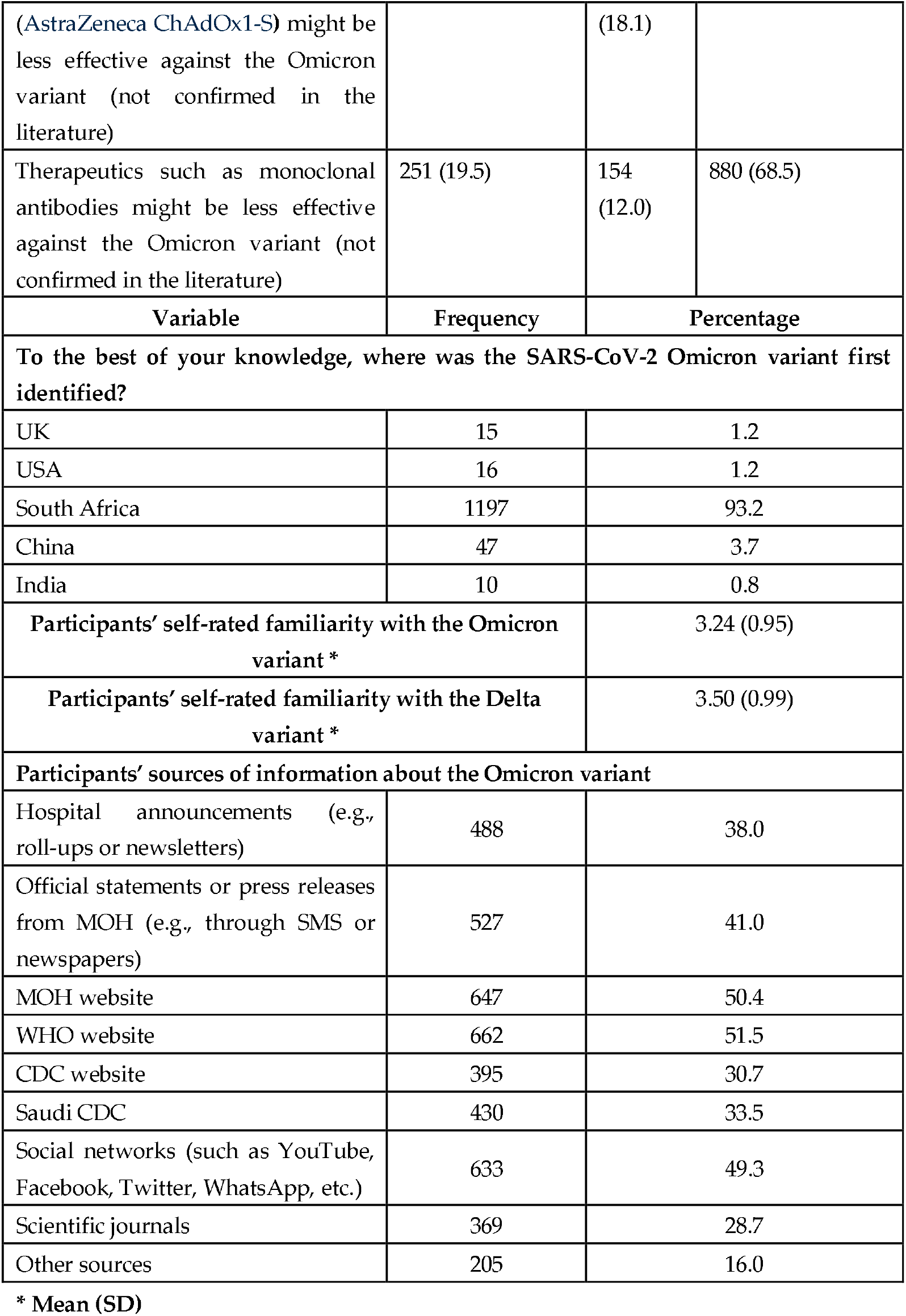
Descriptive analysis of HCWs’ awareness and knowledge of the Omicron variant.

The mean participants’ self-rated familiarity with the Omicron and Delta variants of the SARS-CoV-2 virus was 3.24 (SD 0.95) and 3.5 (SD0.99), respectively, with strong and significant correlation between these scores p < 0.0001 (t = 6.7928, df = 2568). When we explored the participants’ sources of information about the Omicron variant, the MOH and WHO websites were the main sources, but social networks were also used to obtain information in equal measure (about 50%), followed by Official statements (41%), hospital announcements (38%), and the Center of Disease Control (CDC) and Saudi CDC websites, which were accessed almost equally (between 30 and 33%), while scientific journals were lower down on the list (28.7%).

Of the participants, 66.9% agreed with the vaccine’s effectiveness to prevent the spread of Omicron or other future variants, while when considering other preventive measures, such as universal masking, social distancing, and avoiding unnecessary international travel, their agreement was higher (77.8%, 78%, and 71.4%, respectively). Participating HCWs (82.2%) felt that the COVID-19 vaccination should be mandatory for all eligible adult populations, while only 5.1% disagreed (Table 4).

**Table 4:**
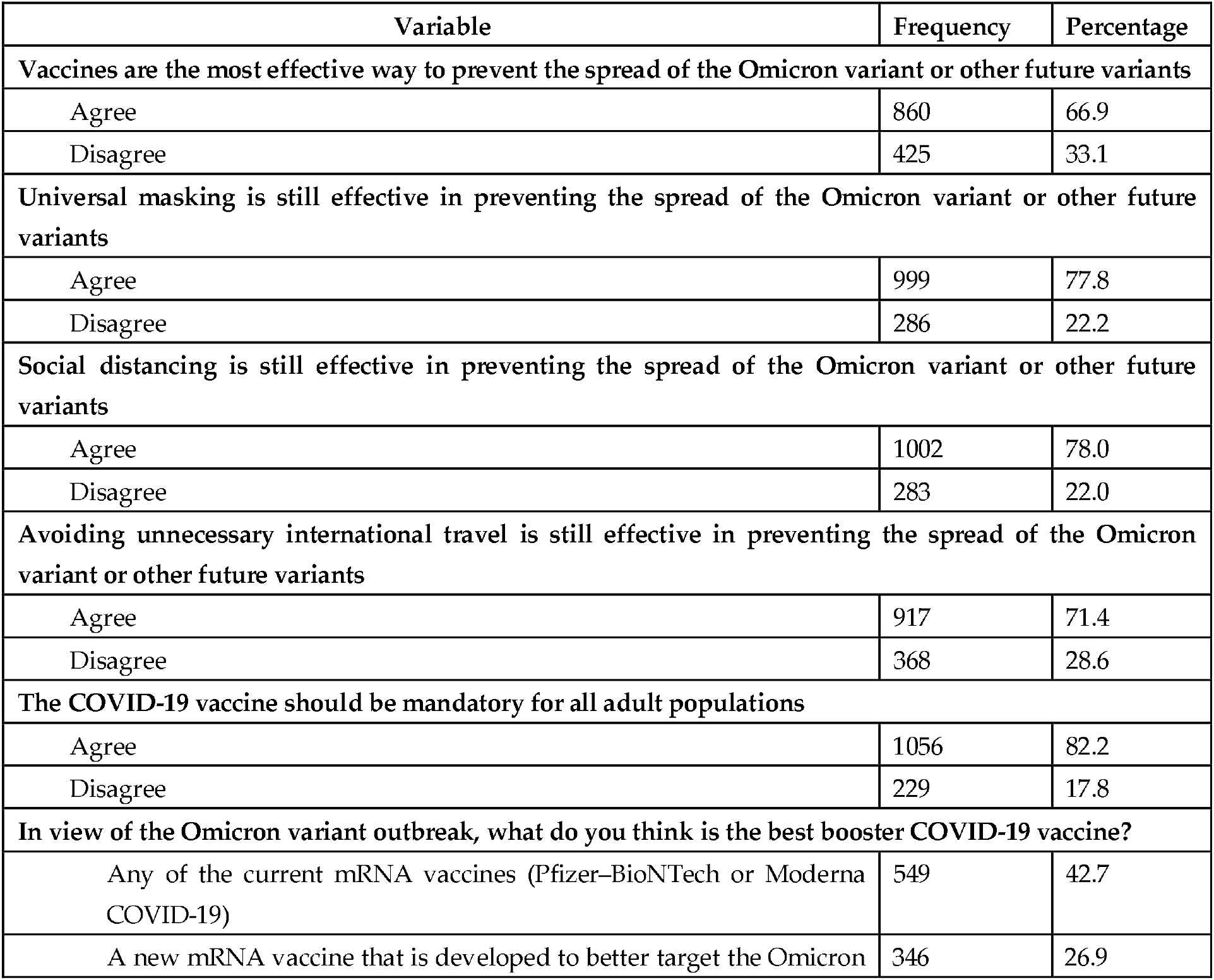
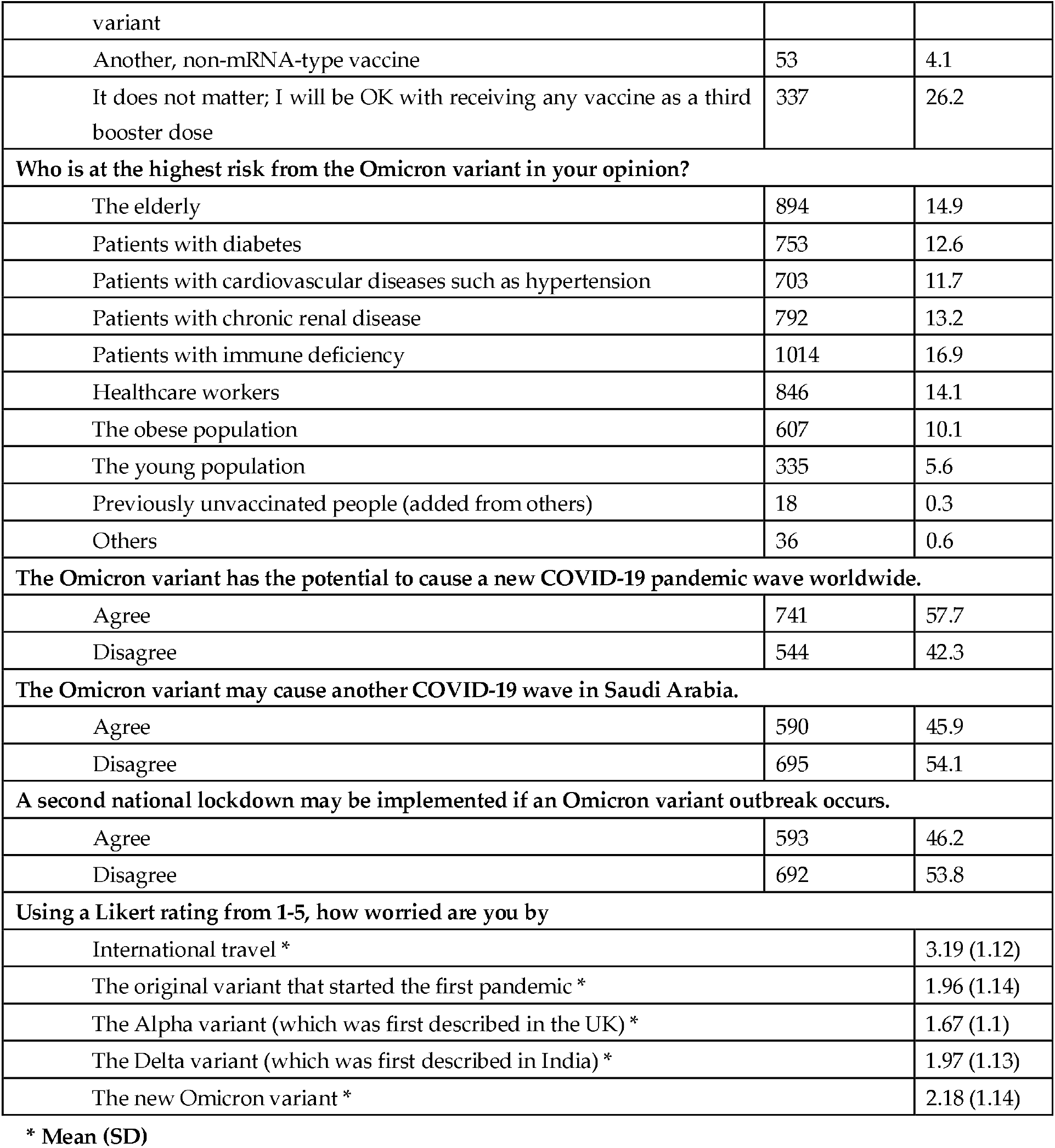
Descriptive analysis of the HCWs’ beliefs, attitudes, and practices concerning the SARS-CoV2 Omicron variant.

Almost half (42.7%) of the HCWs perceived that any of the current mRNA vaccines (Pfizer–BioNTech COVID-19 vaccine or Moderna COVID-19 vaccine) would be the best COVID-19 booster vaccine, while 26.9% felt that a newly developed mRNA vaccine that would better target the Omicron variant is best, and 26.2% agreed to receive any of the available vaccines as a third booster dose (Table 4).

Regarding the participants’ perceived risk from the Omicron variant, 16.9% felt that patients with immune deficiencies are at the highest risk, followed by the elderly (14.9%), HCWs (14.1%), and then patients with chronic medical illnesses (Table 4).

When assessing the participants’ perceptions about future perspectives of the Omicron variant locally and internationally, 57.7% agreed that it has the potential to cause a new COVID-19 pandemic wave worldwide, while only 45.9% agreed that it may cause another COVID-19 wave in Saudi Arabia, and 46.1% agreed that it may cause a lockdown (Table 4). The current cohort of HCWs had the highest worry level in relation to international travel (3.19, SD 1.12), followed by the Omicron variant (2.18, SD 1.14), and their worry levels in relation to the original SARS-CoV-2 and Delta variants were comparable (1.96, SD 1.14 and 1.97 SD, 1.13, respectively), while their worry level in relation to the Alpha variant was the lowest (1.67, SD 1.1).

### HCWs’ perceptions of vaccination and other preventive measures to prevent the spread of SARS-CoV-2 variants

The HCWs’ belief in universal masking, social distancing, and avoiding unnecessary international flights to prevent the spread of SARS-CoV-2 variants correlated strongly and significantly with their belief that vaccines are still the most effective way to prevent the spread of Omicron and other future variants (r = 0.707, 0.675, 0.603, respectively; p < 0.01). Their mean worry level in relation to international travel also correlated significantly with the above measures (r = 0.102, 0.134, 0.157, respectively; p <0.01), which also correlated with their agreement with COVID-19 vaccine mandates for all adults (r = 0.139, p < 0.01). Their mean worry level in relation to the Omicron variant correlated significantly and strongly with their perception of the effectiveness of the above preventive measures, including vaccination to prevent its spread and mandatory vaccination (r = 0.082, 0.103, 0.132, 0.160, 0.114, respectively; p <0.01). Self-rated familiarity with the Omicron or Delta variants correlated strongly and significantly only with their agreement of universal masking as a preventive measure to prevent the spread of other variants, including Omicron, (r = 0.090, 0.094, respectively; p <0.01). Additionally, their familiarity with either variant correlated significantly with their agreement with mandatory vaccination (r = 0.167, 0.142, respectively; p < 0.01) (Table 5).

**Table 5:** Correlation between participants’ perceptions about vaccines, universal masking and social distancing to prevent the spread of the Omicron variant.

| Variable | Vaccines effectiveness<sup>a</sup> | Universal masking effectiveness<sup>b</sup> | Social distancing effectiveness<sup>c</sup> | COVID-19 vaccine mandate<sup>k</sup> |
| --- | --- | --- | --- | --- |
| Universal masking<sup>d</sup> | .707** |  |  |  |
| Social distancing effectiveness<sup>e</sup> | .675** | .889** |  |  |
| Avoiding unnecessary international travel<sup>f</sup> | .603** | .755** | .794** |  |
| Omicron variant potential/new COVID-19 pandemic<sup>g</sup> | -.002 | .011 | .017 |  |
| Omicron variant potential/COVID-19 wave<sup>h</sup> | .005 | -.015 | .000 |  |
| Omicron variant potential/lockdown<sup>i</sup> | .010 | .027 | .051 |  |
| COVID-19 vaccine mandate<sup>k</sup> | .179** | .117** | .113** |  |
| Mean worry level in relation to international travel | .102** | .134** | .157** |  |
| Mean worry level in relation to original variant | .069* | .091** | .111** | .139** |
| Mean worry level in | .065* | .075** | .098** | .148** |

| relation to Alpha variant |  |  |  |  |
| --- | --- | --- | --- | --- |
| Mean in relation to level from Delta variant | .069* | .109** | .124** | .114** |
| Participants' mean worry level in relation to the Omicron variant | .082** | .103** | .132** | .114** |
| Self-rated familiarity with Omicron variant | .090** | .055* | .069* | .167** |
| Self-rated familiarity with the Delta variant | .094** | .062* | .066* | .142** |

a Vaccines are still the most effective way to prevent the spread of Omicron and other future variants. b Universal masking is still effective in preventing variant spread. c Social distancing is still effective in preventing variant spread. d Participants' agreement level with universal masking still being effective in preventing the spread of SARS-CoV-2 variants. e Participants' agreement level with the effectiveness of social distancing to prevent the spread of SARS-CoV-2 variants. f Participants' agreement level with the effectiveness of avoiding unnecessary international travel to prevent the spread of SARS-CoV-2 variants. g Participants' agreement level with the potential of the Omicron variant to cause a new COVID-19 pandemic wave worldwide. h Participants' agreement level with the potential of the Omicron variant to cause another COVID-19 wave in Saudi Arabia. i Participants' agreement level with the potential of the Omicron variant to cause second lockdown. k Participants' agreement level with COVID-19 vaccine mandates.
\*\*. Correlation is significant at the 0.01 level (2-tailed). \* Correlation is significant at the 0.05 level (2-tailed). Note: Correlation (r) coefficients <0.10 are considered non-significant or are weak even if their p values are <0.050.

Multivariate logistic regression analysis of multi-variables related to the HCWs and their perception of the ineffectiveness of the currently available COVID-19 vaccines to prevent the spread of SARS-CoV-2 variants found that the HCWs’ age correlated positively with their agreement with the ineffectiveness of the COVID-19 vaccines when they were aged over 35 years (OR = 1.25, p = 0.235), and significantly when they were aged 55 and above (OR = 1.96, p = 0.034) (Figure 1) (Table 6). HCWs who strongly believed that universal masking and avoiding unnecessary international flights are still effective ways to prevent the spread of mutant variants were in significant disagreement with the ineffectiveness of the vaccines to prevent the spread of current and future mutant variants.

**Figure 1:**
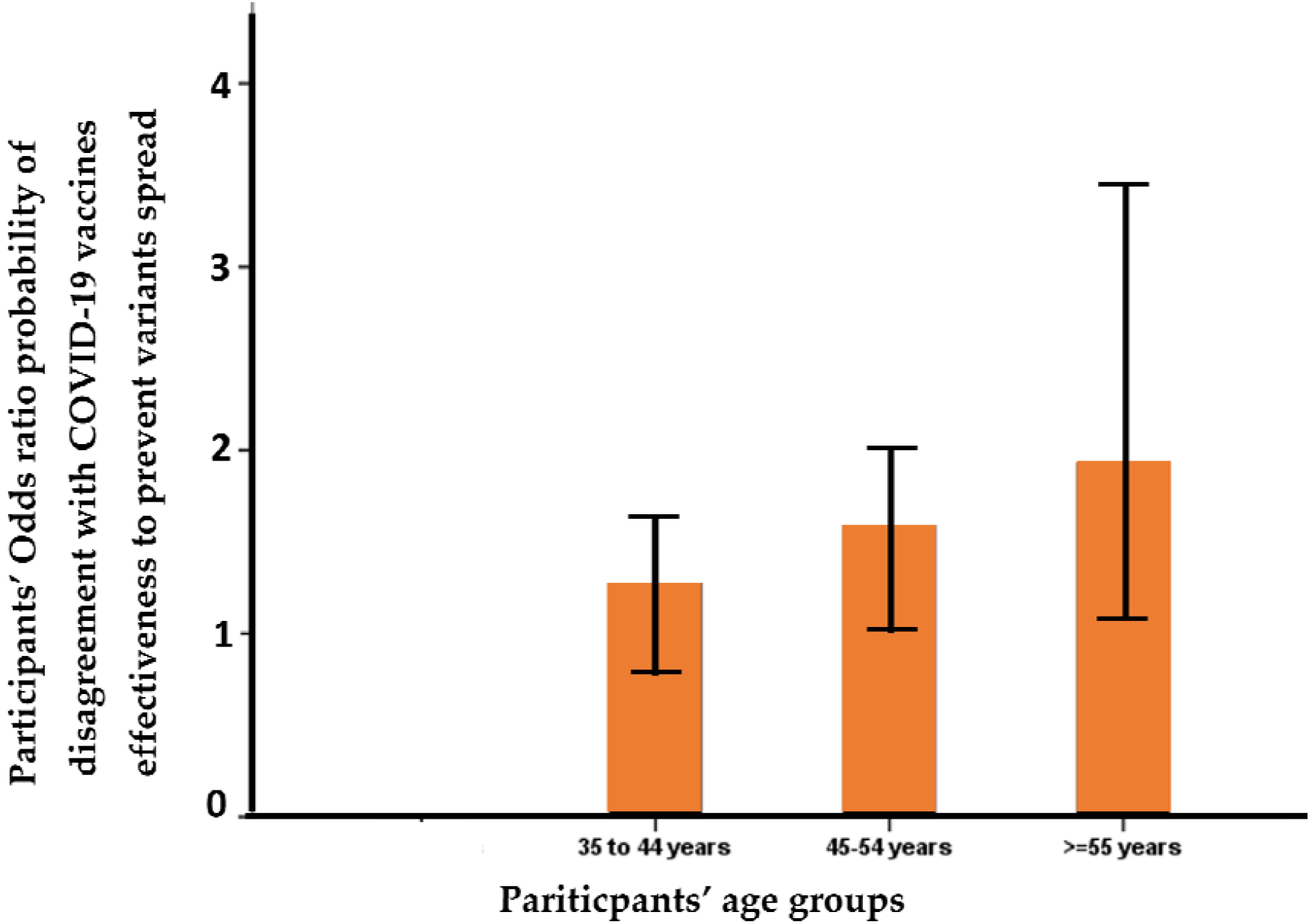
Odds ratio between the participants’ ages and their agreement with the ineffectiveness of the COVID-19 vaccines to prevent the spread of mutant variants; the correlation is significant for those over 55 years old; p = 0.034.

**Table 6:** Multivariate logistic regression analysis of odds of agreement with the ineffectiveness of the COVID-19 vaccines to prevent the spread of mutant variants, especially Omicron.

| Variable | Multivariate<br>adjusted odds<br>ratio | 95% C.I. for OR |  | p value |
| --- | --- | --- | --- | --- |
|  |  | Lower | Upper |  |
| Sex =male | .857 | .609 | 1.208 | 0.379 |
| Age =35 to 44 years | 1.255 | .863 | 1.827 | 0.235 |
| Age =45-54 years | 1.502 | .967 | 2.332 | 0.070 |
| Age >=55 years | 1.965 | 1.054 | 3.663 | 0.034 |
| Work area =ICU | 1.925 | 1.194 | 3.103 | 0.007 |
| Work area = OR | 2.148 | .878 | 5.255 | 0.094 |
| Previously diagnosed with COVID-19 | 1.292 | .897 | 1.863 | 0.169 |
| Does not want to take the COVID-19 vaccine | 1.564 | .833 | 2.937 | 0.164 |
| Believes SARS-CoV-2 variants are expected | .799 | .638 | 1.000 | 0.050 |
| Believes universal masking is still effective at preventing the spread of variants | .318 | .258 | .393 | <0.001 |
| Believes avoiding unnecessary international travel is still effective at preventing the spread of variants | .617 | .499 | .763 | <0.001 |
| Agreement level that the Omicron variant has the potential to cause a new COVID-19 pandemic wave worldwide | 1.208 | .934 | 1.562 | 0.149 |
| Agreement level that COVID-19 vaccination should be mandatory for all adults | .355 | .263 | .478 | <0.001 |
| Source of info: MOH website | .760 | .544 | 1.062 | 0.108 |
| Source of info: WHO website | 1.474 | 1.046 | 2.077 | 0.027 |
| Source of info: CDC website | .768 | .521 | 1.133 | 0.183 |
| Source of info: Scientific journals | .774 | .526 | 1.137 | 0.191 |
| Worry level in relation to the Delta variant | 1.127 | .971 | 1.307 | 0.117 |

Dependent variable: COVID-19 vaccine ineffectiveness to prevent the spread of mutant variants, especially Omicron.

In addition, those who believed that COVID-19 vaccination should be mandatory for all adults or believed that mutant variants are an expected phenomenon also showed strong disagreement with the ineffectiveness of the vaccines. HCWs who worked in critical care areas showed significant agreement with the belief that the COVID-19 vaccines are ineffective in preventing the spread of mutant variants. Regarding the HCWs’ sources of information, those who significantly relied on the WHO did not believe in the efficacy of the vaccines to prevent the spread of current or future mutant variants (Table 6).

An analysis of the characteristics of the surveyed HCWs found that males (compared to females) were in significant agreement with mandatory vaccination of all eligible adults, while age did not correlate with agreement with mandatory vaccination. HCWs who relied on social media as a source of information were in significant agreement with mandatory vaccination for all adults, self-rated familiarity level with the Omicron variant and the agreement that there could be further national lockdowns due to the Omicron variant. Furthermore, the level of agreement with the effectiveness of vaccines to prevent the spread of mutant variants correlated significantly with agreement with mandatory vaccination.

Those who were absolutely unwilling to receive the vaccine were five times more likely to disagree with mandatory vaccination, while those who agreed with the effectiveness of unnecessary travel avoidance to prevent the spread of mutant variants were 1.5 times more likely to disagree with the mandatory vaccination of all eligible adults (Table 7).

**Table 7:**
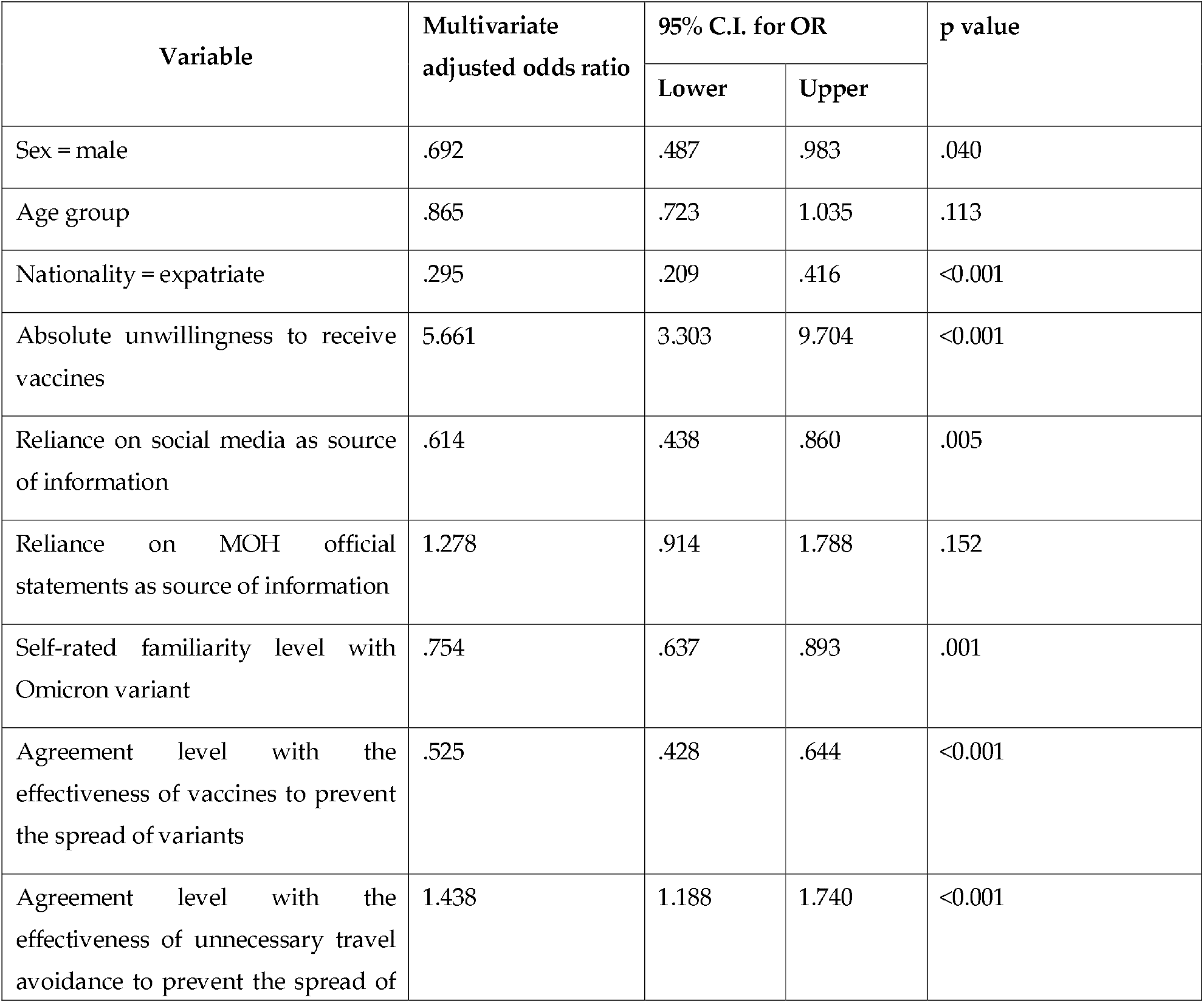
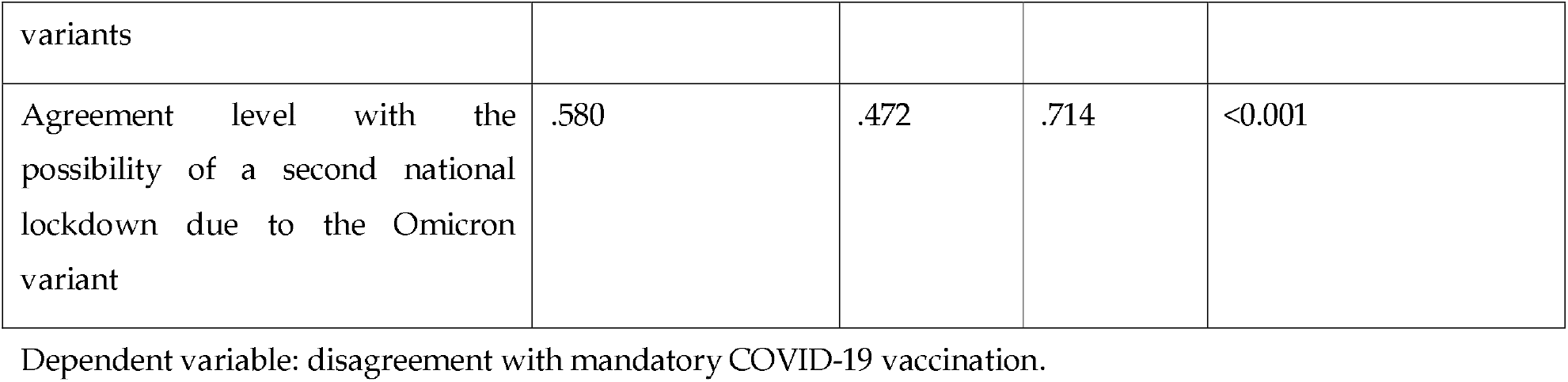
Multivariate logistic regression analysis of odds of disagreement with mandatory COVID-19 vaccination.

## Discussion

This study explored HCWs’ perceptions and awareness of the SARS-CoV-2 Omicron variant after the initial WHO alert, in addition to their agreement with vaccine effectiveness to prevent its spread or other potential future variants. Most (64%) respondents were female, similar to a previous study about Delta variant concerns in KSA. This was also in agreement with other studies showing that the majority of HCWs are female [25-27].

One-fifth of study participants received a booster dose, while the majority reported plans to receive it once they became eligible, with a small percentage (6%) not planning to receive it. The majority (69.6%) in our study chose to receive the booster dose with any of the currently available mRNA vaccines or newly developed ones; this observation mirrors another study conducted in a neighboring country among HCWs, where they also preferred mRNA vaccines[28]. Such behavior could be partly due to the hematological side effects that have been recorded with the viral vector vaccines [28]. In addition, this could signal vaccine selectivity (VS), which is currently an emerging public health challenge due to the multiplicity of vaccine options. Another explanation is vaccine hesitancy (VH), which could be related to reliance on social media as the main source of information, which was reported by almost half of the participants, as opposed to less than one-third who reported utilizing scientific journals as a main source of information; therefore, increased access to peer-reviewed journals should be strongly encouraged among HCWs[25]. Keeping in mind the double edge sword effects the social media plays on individuals’ decision making, as social media spreads information that’s not scientifically always verified, puts their personal touch or psycho-cognitive flavor that drives the audience, therefore our observation of social media being highly used source of information by the surveyed HCWs can explain some observations like (VS), (VH) and HCWs significant agreement with mandatory vaccination for all adults. Such agreement might be a reflection of their self-rated familiarity level with the Omicron variant (stemming from the information they acquired from social media) that also correlated significantly with their agreement with mandatory vaccination. Social media spread with the emergence of the Omicron variant a huge load of information that was potentially driving all sectors of the society to abide with all the measures of protection including vaccination, from SARS-CoV-2 including its new Omicron variant.

The primary concerns regarding Omicron are whether it is more contagious, more virulent, or both, in comparison to other variants, and how likely it would be to evade natural (post previous COVID-19 infection) and vaccine-induced immunity [11,29-31]. However, with more than 30 mutations and some deletions that are similar to other variants, increased transmissibility and higher antibody escape were expected [32]. However, with few clinical and epidemiological data to help define the true threat of Omicron, participants’ mixed knowledge responses are expected, as has been observed in our study, in which they even extended to their belief in the effectiveness of the vaccines to control its spread [33]

Among our study participants, regarding the perceived effectiveness of various methods to control the spread of the Omicron variant, social distancing and universal masking scored the highest, followed by avoiding international flights and, lastly, vaccination. Again, such observations shed light on their questioning of the vaccine’s effectiveness against Omicron due to its many mutations, especially involving the spike protein. Their perceived effectiveness of the current COVID-19 vaccines against the Omicron variant was 67%. This represents a decline in HCWs’ perception of vaccine efficacy compared to our earlier study, performed in December 2020 during the initial rollout of the COVID-19 vaccines, which was 80.2% [34]. Such findings reflect declining confidence in vaccine efficacy against emerging variants, which is noteworthy since their concerns about efficacy have been stated as one of the main reasons for vaccine hesitancy [35]. COVID-19 vaccine-related uncertainty and challenges persist in the face of combacting emerging SARS-CoV-2 variants. Immune escape is a real concern with the Omicron variant. According to findings from South Africa, the efficacy of the Pfizer-BioNTech vaccine against the Omicron variant was significantly reduced, with a 41-fold lower level of neutralizing antibodies when compared to the wild-type variant of SARS-CoV-2 [10].

The fact that 82.2% of HCWs agree with mandatory COVID-19 vaccination for all adults is an interesting finding. Mandating immunizations for HCWs or other business employees is not only ethical, but also legal. Employers have the right to require vaccination as a condition of employment, as the Equal Employment Opportunity Commission and courts have stated [36]and a multi-society statement on COVID-19 vaccination had been also released[37]. This right applies to both vaccinations approved for emergency use and those that have been fully approved by the FDA. Mandates, however, can erode public support, causing a backlash and even lowering vaccine uptake. There are no established processes to enforce population-wide vaccination obligations in most countries, despite the fact that employers, healthcare providers, and educational institutions can monitor compliance with mandates. Requiring COVID-19 immunizations for HCWs, for example, is not a new practice; rather, it is a continuation of a long-standing policy. Vaccination against influenza, hepatitis B, and other infectious diseases has long been mandatory in many health-care settings [38]. Whether to mandate COVID-19 vaccination for HCWs and/or all employees and how to implement such a policy is ultimately a local authority decision. For instance, the local authorities in both New York and California announced a vaccine mandate for state employees [39].

We observed that HCWs’ beliefs in infection control measures that have been mandated and optimized during the COVID-19 pandemic namely, universal masking, social distancing, and vaccination as preventive methods for the spread of the Omicron variant or other future variants [40,41] all correlated positively and significantly with their belief in mandatory vaccination for all eligible adults. They still believed that avoiding unnecessary international travel is an effective measure in preventing the spread of SARS-CoV-2 variants, even after widespread vaccination, perhaps due to the emergence of at least three variants, one of them with high transmissibility and serious clinical morbidity and high mortality (the Delta variant). However, there is a huge consequence of travel ban and lockdown with any emerging variant[42,43].

HCWs who believed that SARS-CoV-2 mutations are expected and will reoccur were significantly less in agreement with the belief that the vaccines are ineffective against the spread of the Omicron variant or other potential future variants, which is an expected and healthy belief, as the best preventive measure to contain the pandemic at present is widespread vaccination This has been observed in HCWs’ stringent behavior of abiding by the infection prevention measures, such as universal masking and social distancing, for the same purpose [40,41], which has also been observed in our results, in which agreement with the effectiveness of vaccination to prevent the spread of mutant variants is significantly and positively associated with agreement with mandatory vaccination.

HCWs in critical care areas believed significantly in the ineffectiveness of the vaccines to prevent the spread of mutant variants, including Omicron. This might be explained by the multiple mutations that the Omicron variant has and the critical care workers’ anxiety regarding the current vaccine’s efficacy against it, being frontliners exposed to severe cases. In the same vein, we observed that HCWs who relied on the WHO as a main source of information disagreed with the vaccine’s effectiveness to prevent the spread of the mutant strains, including Omicron, as it is a scientific source that releases only evidence-based information, which has not thus far proven the vaccine’s efficacy in that regard.

Male gender was significantly associated with agreement with mandatory vaccination of all eligible adults. This attitude has been observed in multiple studies, as males were more inclined toward vaccine uptake and had much less vaccine hesitancy, while age did not correlate with mandatory vaccination agreement [28,44].

The level of agreement with the effectiveness of vaccines to prevent the spread of mutant variants was significantly associated with agreement with mandatory vaccination, and the same was observed with the level of agreement with the belief that there might be a second national lockdown due to the Omicron variant. This behavior from HCWs—of stringent abidance with vaccination—correlates with their expectation of its effectiveness to prevent the spread of any mutant variant, and their fear of a national lockdown, not unexpectedly, drove them to have an unwavering belief in mandatory vaccination as the best modality to prevent it. However, they did not perceive the effectiveness of unnecessary travel avoidance to be effective in preventing the spread of mutant variants or being superior to mandatory vaccination. Early lockdown and social distancing in the initial phase of the pandemic were useful in slowing the spread of SARS-CoV-2 in Saudi Arabia[45]. In addition, during the peak of the initial lockdown the level of anxiety was associated with being with family members at risk of SARS-CoV-2 infection. Thus, it is understandable that HCWs fear the occurrence of second lockdown with the emergence of the Omicron variant[46].

### 3.1 Study limitations and strengths

While this research is subject to the limitations of cross-sectional studies, such as sampling, response rate, and uneven geographical representation, the fact that we carried out the study early on in the spread of the Omicron variant also limited our ability to assess the awareness of the HCWs in relation to the variant, especially given the scarce scientific data available after it was first identified. Despite this, the scientific evidence regarding the current vaccines’ effectiveness against the Omicron variant will take a significant amount of time to materialize. As with the evolving situation in regard to variants, HCWs’ experiences and perceptions are also likely to change. Moreover, HCWs’ experiences may differ from one setting to another.

## 4. Conclusion

This is the first national survey in Saudi Arabia that was conducted in the first week of the WHO Omicron variant announcement, to address HCWs’ awareness of the variant, acceptance of vaccine booster doses and agreement with vaccination to prevent the spread of SARS-CoV-2 Omicron variant or future variants. Only about two-thirds of HCWs perceived that vaccination is the best option to prevent the spread of the Omicron variant, indicating the need for future studies to explore the performance of currently available vaccines in terms of the protection they offer against the current variants. At the same time, the HCWs had a strong belief in preventive measures, such as universal masking and social distancing, to prevent the spread of Omicron or future variants, which should still be maintained in practice, at least in healthcare institutions, to prevent nosocomial spread until further evidence can be elucidated.

## Data Availability

All data produced in the present study are available upon reasonable request to the corresponding author.

## Abbreviations

CDC: Center for Disease Control and Prevention
COVID-19: Coronavirus disease 2019
SARS-CoV-2: Severe acute respiratory syndrome coronavirus 2
WHO: World Health Organization

## Conflicts of interest

None declared.

## Ethics approval and consent to participate

The study was approved by the institutional review board of King Saud University (approval 21/01039/IRB).

## Acknowledgments

This research has been financially supported by Prince Abdullah Ben Khalid Celiac Disease Research Chair, under the Vice Deanship of Research Chairs, King Saud University, Riyadh, Kingdom of Saudi Arabia. The research team is thankful for the statistical data analysis consultation offered by www.hodhodata.com.

## Author contributions

M.H.T., S. Alenezi, M.A., F.A., K.A., F.A., S. Al-Subaie, M.B., Z.M., and J.A. conceptualized the study, analyzed the data, and wrote the manuscript. R.A., R.B., F.A., A. Alrabiaah, A. Alhaboob, A. Alaraj, N.S.A., R.H., L.A., N.A., A.J., A. Aleyadhy, and W.A. contributed to the study design; collected, analyzed, interpreted data; and edited the manuscript.

All authors reviewed and approved the final version of the manuscript.

## Notes

### Competing Interest Statement

The authors have declared no competing interest.

### Funding Statement

This study did not receive any funding.

### Author Declarations

Ethics committee/IRB of King Saud University gave ethical approval for this work (approval 21/01039/IRB).

### Summary of Updates

Updated Discussion with more peer-reviewed publications.

